# Effectiveness of Subcutaneous Casirivimab and Imdevimab Relative to no COVID-19 Antibody Treatment Among Patients Diagnosed With COVID-19 in the Ambulatory Setting

**DOI:** 10.1101/2022.06.20.22276636

**Authors:** Jessica J. Jalbert, Mohamed Hussein, Vera Mastey, Robert J. Sanchez, Degang Wang, Dana Murdock, Laura Farinas, Jonathan Bussey, Carlos Duart, Boaz Hirshberg, David M. Weinreich, Wenhui Wei

**Affiliations:** Regeneron Pharmaceuticals, Tarrytown, NY; CDR Maguire, Tallahassee, FL

## Abstract

**Importance:** Data on real-world effectiveness of subcutaneous (SC) administration of casirivimab and imdevimab (CAS+IMD) for treatment of COVID-19 are limited.

**Objective:** To assess effectiveness of SC CAS+IMD vs no COVID-19 antibody treatment among patients diagnosed with COVID-19 in ambulatory settings during the Delta-dominant period prior to Omicron emergence.

**Design:** Retrospective cohort study.

**Setting:** Encrypted linked data between Komodo Health closed claims database and CDR Maguire Health & Medical database.

**Participants:** Patients with COVID-19 in ambulatory settings between August 1, 2021 and October 30, 2021 treated with SC CAS+IMD were exact- and propensity score-matched to up to 5 untreated patients who were treatment-eligible under the Emergency Use Authorization (EUA)

**Exposure:** Subcutaneous CAS+IMD.

**Main Outcomes and Measures:** Composite endpoint of 30-day all-cause mortality or COVID- 19-related hospitalization. Kaplan-Meier estimators were used to calculate composite risk overall and across subgroups including age, COVID-19 vaccination status, immunocompromised, and elevated risk defined as age ≥ 65 years or 55-64 years with body mass index ≥ 35 kg/m^2^, type 2 diabetes, chronic obstructive pulmonary disease, or chronic kidney disease. Cox proportional-hazards models were used to estimate adjusted hazard ratios (aHR) and 95% confidence intervals (CI).

**Results:** Among 13 522 patients treated with SC CAS+IMD, 12 972 (95.9%) were matched to 41 848 EUA-eligible untreated patients; patients were 57-58% female, with mean age between 50 and 52 years. The 30-day composite outcome risk was 1.9% (95% CI, 1.7-2.2; 247 events) and 4.4% (95% CI, 4.2-4.6; 1822 events) in the CAS+IMD-treated and untreated cohorts, respectively; CAS+IMD treatment was associated with a 49% lower risk (aHR 0.51; 95% CI, 0.46-0.58). Treatment was also associated with a 67% lower 30-day risk of all-cause mortality (aHR 0.33, 95% CI, 0.18-0.60). Treatment effectiveness was consistent regardless of vaccination status and across subgroups, including those at elevated risk (aHR 0.51, 95% CI 0.42-0.60) or immunocompromised (aHR 0.34, 95% CI 0.17-0.66).

**Conclusions and Relevance:** Subcutaneous treatment with CAS+IMD is effective for reducing all-cause mortality or COVID-19-related hospitalization in patients diagnosed with COVID-19 and managed in real-world outpatient settings during the Delta-dominant period. Effectiveness is maintained among immunocompromised, vaccinated, and elevated risk patients.

## Introduction

While vaccines remain the primary strategy for control of the coronavirus 2019 (COVID-19) pandemic caused by severe acute respiratory syndrome coronavirus 2 (SARS-CoV-2), they require development of active immunity to COVID-19 over time. In contrast, neutralizing monoclonal antibodies (mAbs) against the SARS-CoV-2 spike protein confer immediate passive immunity for SARS-CoV-2 variants that remain sensitive to the mAbs and can be used, for pre- or post-exposure prophylaxis or early treatment.^1–7^

In a clinical trial, intravenous (IV) administration of mAbs casirivimab and imdevimab (CAS+IMD; Regeneron Pharmaceuticals, Inc.) was associated with a 71% reduction in all-cause mortality or COVID-19-related hospitalization in patients diagnosed with COVID-19 in ambulatory settings.^2^ These mAbs were granted Emergency Use Authorization (EUA) by the US Food and Drug Administration (FDA) for treatment of non-hospitalized COVID-19 patients who are at risk for severe disease first as IV administration in November 2020 and in June 2021, as subcutaneous (SC) administration when IV infusion is not feasible or would lead to delay in treatment.^8^ In January 2022, with the surge in the Omicron variant (B.1.1.529), the FDA amended the EUA for CAS+IMD to exclude its use in geographic regions where infection or exposure is likely due to to a variant not susceptible to the treatment.^9^ Consequently, it is not currently authorized for use in the US.

While the majority of real-world studies assessing the effectiveness of CAS+IMD reported 50%- 78% reductions in the risk of hospitalization,^10–23^ the effectiveness of SC CAS+IMD was not specifically assessed. A study that did evaluate SC CAS+IMD reported that treated patients were 56% less likely to be hospitalized or die than untreated patients.^24^ However, the results reflect the experience at one medical center between July and October 2021 in 652 patients.

In June 2021, the Florida Department of Health and Florida Division of Emergency Management deployed COVID-19 mAb therapy treatment sites in Florida. A health disaster management company, CDR Maguire Health & Medical (“CDR Health”), was commissioned to manage the COVID-19 public health crisis. Between August and November 2021, CDR Health treated ∼115 000 patients with SC CAS+IMD. The objective of this study was to compare the effectiveness of SC CAS+IMD to no COVID-19 mAb treatment on 30-day all-cause mortality or COVID-19-related hospitalization among patients diagnosed with COVID-19 in the ambulatory setting who were eligible to receive treatment under the EUA.

## Methods

### Data sources

We conducted a retrospective cohort study using closed administrative claims data from the Komodo Health claims database.^25^ As there is no code for distinguishing between SC and IV administration of CAS+IMD in administrative claims data, the Komodo Health data were linked to data from CDR Health, which administered only SC CAS+IMD. The database linkage used Datavant’s encryption and tokenization technology (San Francisco, CA; https://datavant.com)^26^

The dataset resulting from the linkage was used to construct the treatment arm and to identify a control group of patients not treated with COVID-19 mAbs. In addition to date of SC CAS+IMD administration, CDR Health data were used to ensure that patients in the untreated control group had not been treated with SC CAS+IMD.

Since all data are de-identified and fully compliant with HIPAA, institutional review board/ethics committee approval was not required.

### Study Population

The treated cohort consisted of patients treated with SC CAS+IMD between August 1, 2021 and October 30, 2021 (index date = date of CAS+IMD administration) who had not received other COVID-19 mAbs (bamlanivimab monotherapy, bamlanivimab and etesevimab, or sotrovimab) within 6 months prior to or on the index date. This period is concurrent with when the Delta variant (B.1.617.2) became dominant in the US,^27^ and prior to the spread of Omicron.^28^ Patients in the untreated cohort were those diagnosed with COVID-19 (ICD-10: U07.1) during the same period and not treated with COVID-19 mAbs. Given that ∼90% of SC CAS+IMD-treated patients did not have a documented COVID-19 diagnosis in the 10 days prior to treatment administration in the Komodo claims data, an index date could not be assigned to the untreated patients based on the distribution of days between diagnosis and treatment as was done previously.^29^ For untreated patients, the index date was the COVID-19 diagnosis date; if multiple COVID-19 diagnoses were available, the first diagnosis was selected, and to identify incident COVID-19 diagnoses, patients were required to have no evidence of a prior COVID-19 diagnosis within 30 days pre-index. This approach excludes the immortal-time, which would have favored treatment had follow-up started for both groups on the COVID-19 diagnosis date, but introduces survival bias that was adjusted for analytically.^30^

Patients in both cohorts were also required to be ≥12 years old on the index date, be continuously enrolled in medical and pharmacy benefits for ≥6 months pre-index (ie, baseline), meet EUA criteria for CAS+IMD treatment at index,^8^ and have a valid date of death if deceased. Patients who were hospitalized or dead on the index date were excluded.

### Outcomes

The primary endpoint was the composite outcome of 30-day all-cause mortality or COVID-19- related hospitalization, and a secondary endpoint was 30-day all-cause mortality. Komodo uses Social Security Administration (SSA) data, a private obituary data source, and a private claims mortality dataset to identify mortality. COVID-19-related-hospitalization was defined as a COVID-19 diagnosis as the primary or admitting diagnosis. Patients were followed from the index date until the occurrence of the outcome or a censoring event, which included receipt of another COVID-19 mAb, the end of 30-day risk period, healthcare plan disenrollment, or study end date.

### Study variables

Baseline demographic variables included age as a continuous variable and categorized by age groups (12-17, 18-54, 55-64, ≥65 years), sex, and geographic region (Florida resident vs not). The Charlson Comorbidity Index (CCI)^31^ was derived by identifying the presence of comorbidities over the baseline period. In addition to body mass index (BMI), which was categorized as not overweight, overweight, obese, severely obese, morbidly obese, and missing, specific comorbidities included cardiovascular disease (myocardial infarction, hypertension, atrial fibrillation, heart failure; ischemic/hemorrhagic stroke, or venous thromboembolism); chronic respiratory disease (asthma, chronic obstructive pulmonary disease, emphysema, obstructive sleep apnea, pulmonary fibrosis, or cystic fibrosis); chronic kidney disease stage 3-5 or renal failure requiring dialysis; B-cell deficiency (primary, secondary, or drug-induced; eTable 1); and diabetes (type 1 or 2). The occurrence of ≥1 hospitalization and emergency room/urgent care visit for any reason during the baseline period was also captured.

The following risk factors for use of CAS+IMD under the EUA were identified during the baseline period up to and including the index date: age ≥65 years on index date; age 12-17 years on index date with BMI ≥85th percentile for age and sex based on CDC growth charts^32^; BMI >25 kg/m^2^; pregnancy; diabetes; chronic lung disease; immunosuppressive disease; history of immunosuppressive treatment; cardiovascular disease, hypertension, or congenital heart disease; sickle cell disease; and neurodevelopmental disorders. Since EUA risk factors of chronic kidney disease, cardiovascular disease or hypertension, and use of medical-related technological dependence could be an outcome of COVID-19 infection, their presence was assessed over the baseline period only.

### Statistical analysis

#### Matching

To derive adjusted estimates, both exact and probabilistic matching methods were used. Propensity scores (PS), derived using logistic regression, predicted the probability of CAS+IMD treatment vs no treatment given age (as a continuous variable), sex,index month, 3-digit zip code (Florida residents) or state (non-Florida residents), individual EUA criteria, BMI category, CCI score (as a continuous variable), prior COVID-19 vaccination (≥1 COVID-19 vaccine during baseline), and baseline healthcare resource utilization (prior all-cause hospitalization and all-cause emergency room/urgent care visits). Each treated patient was matched to up to 5 untreated patients on a caliper width of 0.2 of the standard deviation of the logit of the estimated PS^33^ and exact-matched on index month and 3-digit zip code (Florida residents) or state (non-Florida residents). Standardized mean differences (SMD) were used to assess balance between groups; SMD >0.1 indicated imbalance and unbalanced variables were included directly in the outcome model.^34^

#### Primary analysis

Baseline characteristics were reported for SC CAS+IMD and untreated EUA-eligible patients using means (SD) and medians (Q1-Q3) for continuous variables, and number and frequency for categorical variables. Kaplan-Meier estimators were used to estimate the 30-day risk of the composite outcome and mortality among the matched patients^35^ with 95% confidence bands constructed using the Hall-Wellner method^36^ and log-rank tests used to compare survival distributions.

Adjusted hazard ratios (aHR) with 95% confidence intervals (CI) were derived using Cox proportional-hazard models that fit the model to the matched pairs, and used robust sandwich variance estimators to account for clustering within matched sets.^37^ Since starting follow-up on the date of treatment for treated patients and the COVID-19 diagnosis for untreated patients excludes immortal-time and can introduce a survival bias, a correction factor was derived to account for this bias.^30^ This bias was removed by dividing the aHR as well as the 95% upper and lower CI values by the correction factor T_0_/T_0_+T_IT;_ T_IT_ denotes the observed immortal person-time which, in our study, is the number of days between the COVID-19 diagnosis and the date of treatment among treated patients, and T0 is person-time in the untreated group. The correction factor was derived assuming that T_IT_ was at most 10 days (the maximum number of permissible days between symptom onset and treatment, per the EUA) but could be shorter if the patient experienced the outcome or was censored. The actual number of days between the COVID-19 diagnosis and treatment date was used for matched treated patients with a non-missing COVID- 19 diagnosis date. For T0, matched EUA-eligible untreated patients contributed up to 30 days, the occurrence of the event, or censoring, whichever came first relative to T_0_. The survival bias was then accounted for by dividing aHR and 95% CIs by the correction factor.

#### Subgroup analyses

Subgroups of interest included age stratified as 12-17, 18-54, 55-64, and ≥65 years; elevated risk defined as age ≥65 years or 55-64 years with BMI ≥35 kg/m^2^, type 2 diabetes, chronic obstructive pulmonary disease, or chronic kidney disease; immunocompromised patients, overall and by type of B-cell deficiency (primary, secondary, or drug-induced); and prior COVID-19 vaccination. The same matched set of patients was used to evaluate effectiveness across subgroups. Kaplan-Meier estimators were used to determine 30-day outcome risks for subgroups. Adjusted HRs comparing treated versus untreated cohorts across subgroups were derived using Cox proportional-hazard models and including the interaction term between treatment and the subgroup. The adjusted estimates of subgroups were derived using the same matched set of patients as the primary endpoint and accounted for clustering of patients. No adjustments were made for multiplicity. As in the primary analysis, a correction factor was applied to aHRs and 95% CIs to account for potential survival bias.

#### Sensitivity analyses

Sensitivity analyses conducted to assess robustness of the results included: 1) deriving the correction factor using the subset of matched SC CAS+IMD-treated patients with a non-missing COVID-19 diagnosis date in the Komodo claim database; 2) modifying the definition of COVID-19-related hospitalization to COVID-19 as the primary diagnosis only; 3) modifying inclusion/exclusion criteria so that only the untreated patients were required to meet the EUA criteria on the index date; and 4) requiring 3 months (versus 6 months) of continuous healthcare plan enrollment pre-index.

The analytic file was created and all analyses were conducted using SAS Version 9.4 (SAS Institute, Cary, NC).

## Results

### Linkage and Cohort Creation

Among 90 133 patients treated with SC CAS+IMD in the CDR Health database, 79 295 patients (88.0%) were successfully linked to the Komodo claims and 13 522 of these were eligible for matching (eFigure 1). After applying inclusion/exclusion criteria to 5 132 798 patients in the Komodo database with a COVID-19 diagnosis during the study period, 828 087 patients were eligible for matching (eFigure 1). A total of 12 972 of the 13 522 (95.9%) SC CAS+IMD-treated patients were exact- and PS-matched to 41 848 EUA-eligible untreated patients.

**Figure 1.**
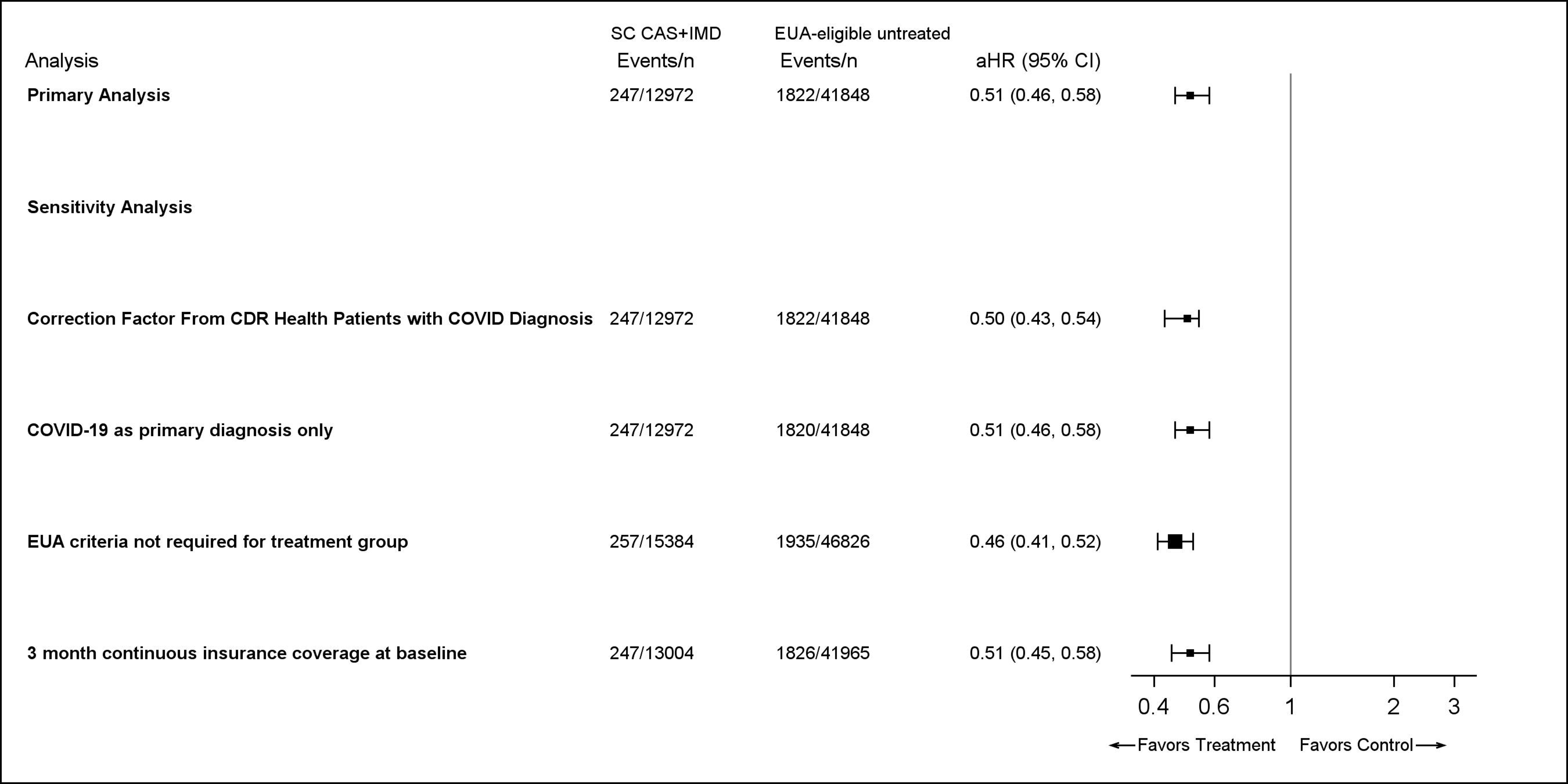
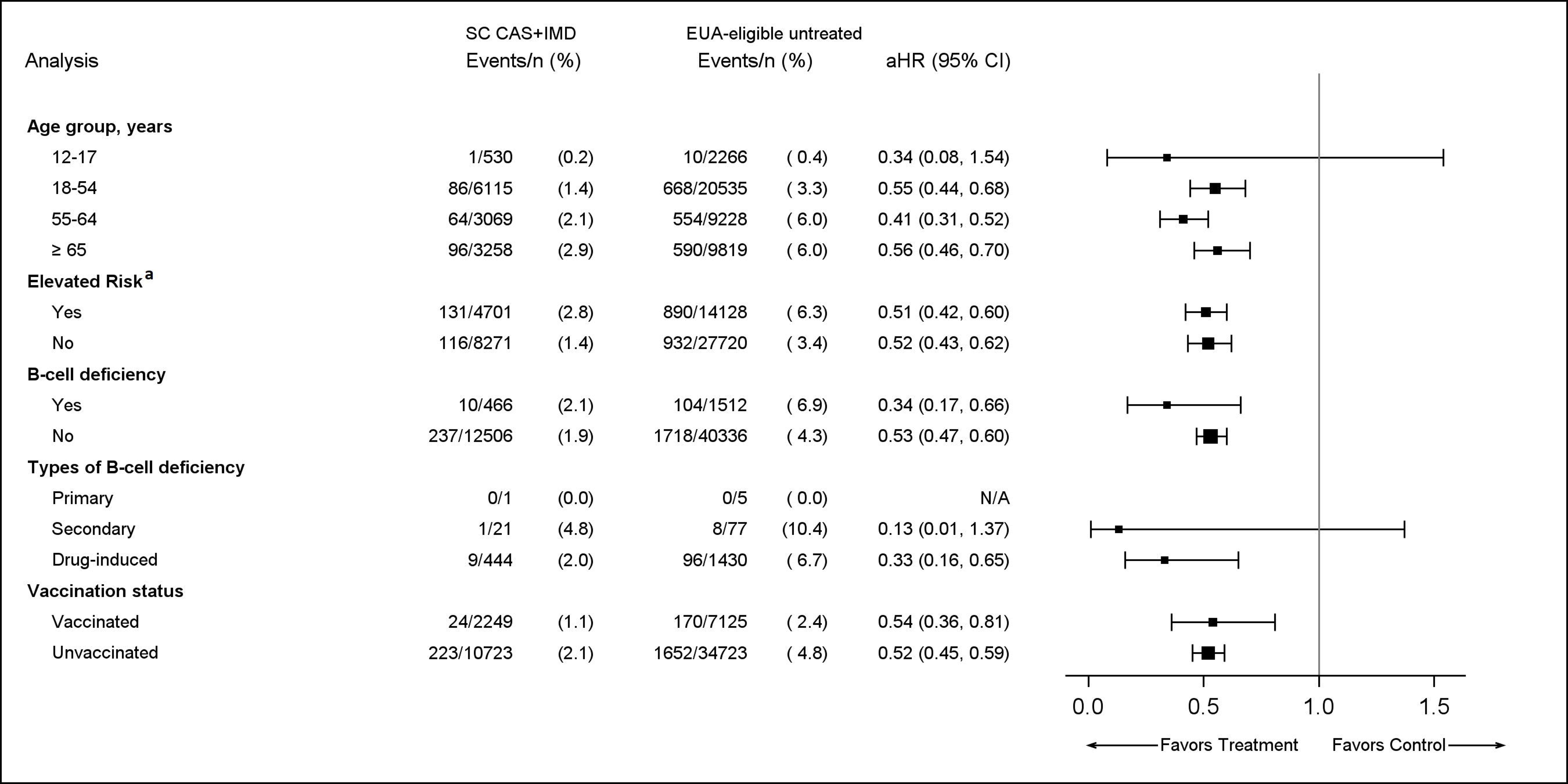
Kaplan-Meier Curves for 30-Day Outcomes Among Patients Diagnosed With COVID-19 in the Outpatient Setting **A) Composite outcome of all-cause mortality or COVID-19-related hospitalization B) All-cause mortality** CAS+IMD indicates casirivimab and imdevimab; EUA, Emergency Use Authorization; SC, subcutaneous.

Prior to matching, cohorts were imbalanced on several variables; notably, the treated cohort was older with a higher proportion at elevated risk (eTable 2). After matching, per the SMDs, all variables except region and index month were balanced (Table 1). The matched populations were 57-58% female), with mean age between 50 and 52 years and 34-36% at elevated risk (Table 1).

**Table 1.**
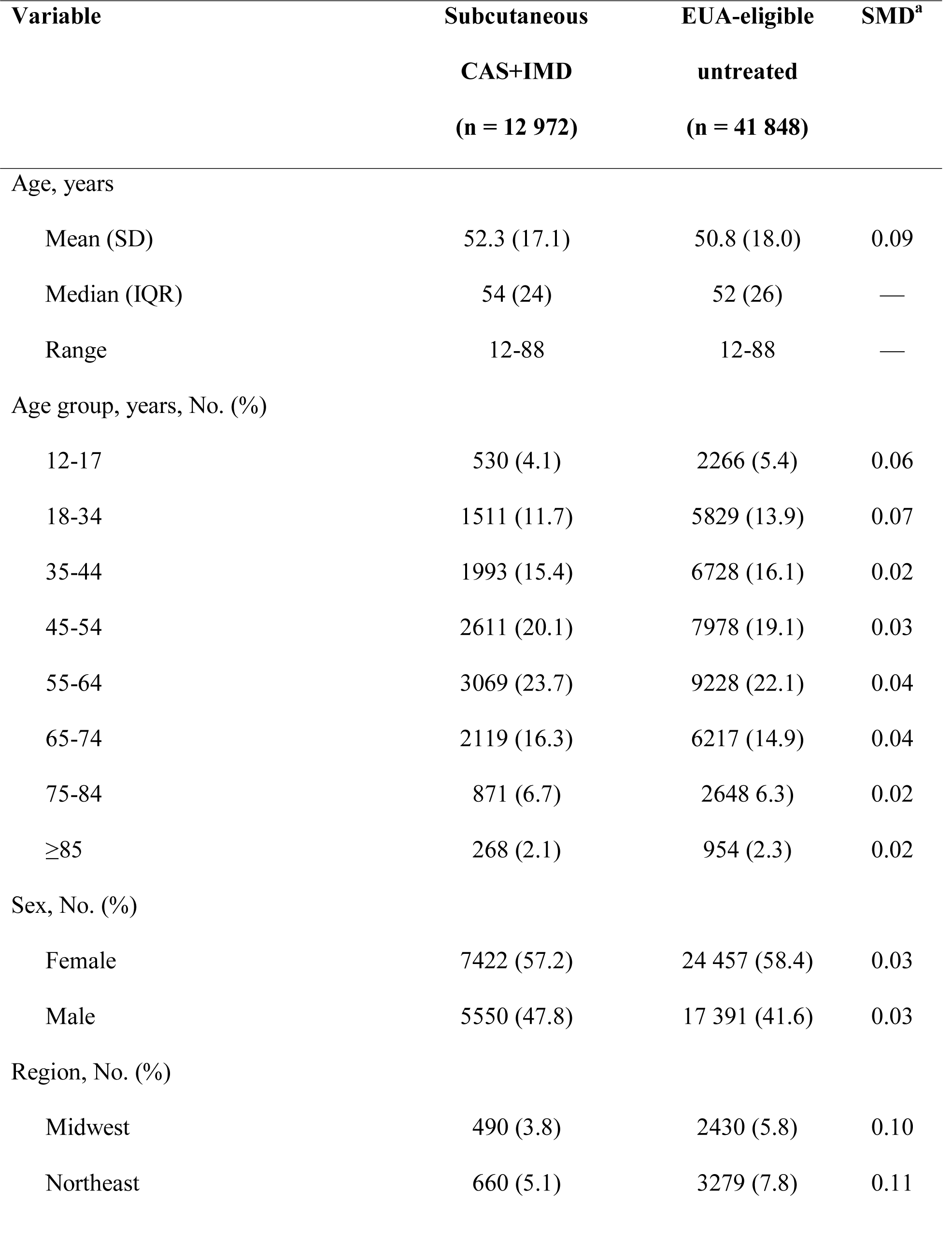
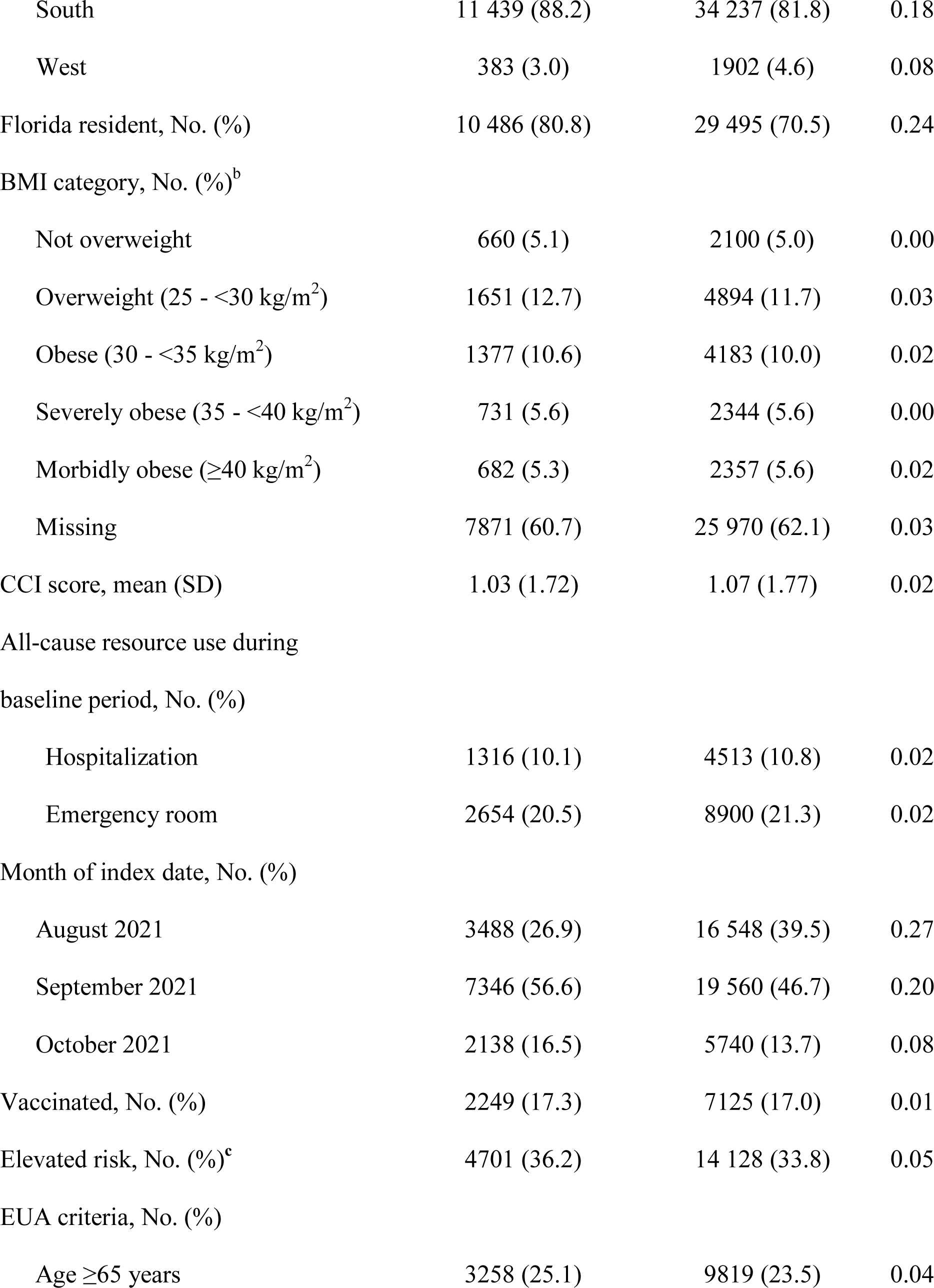
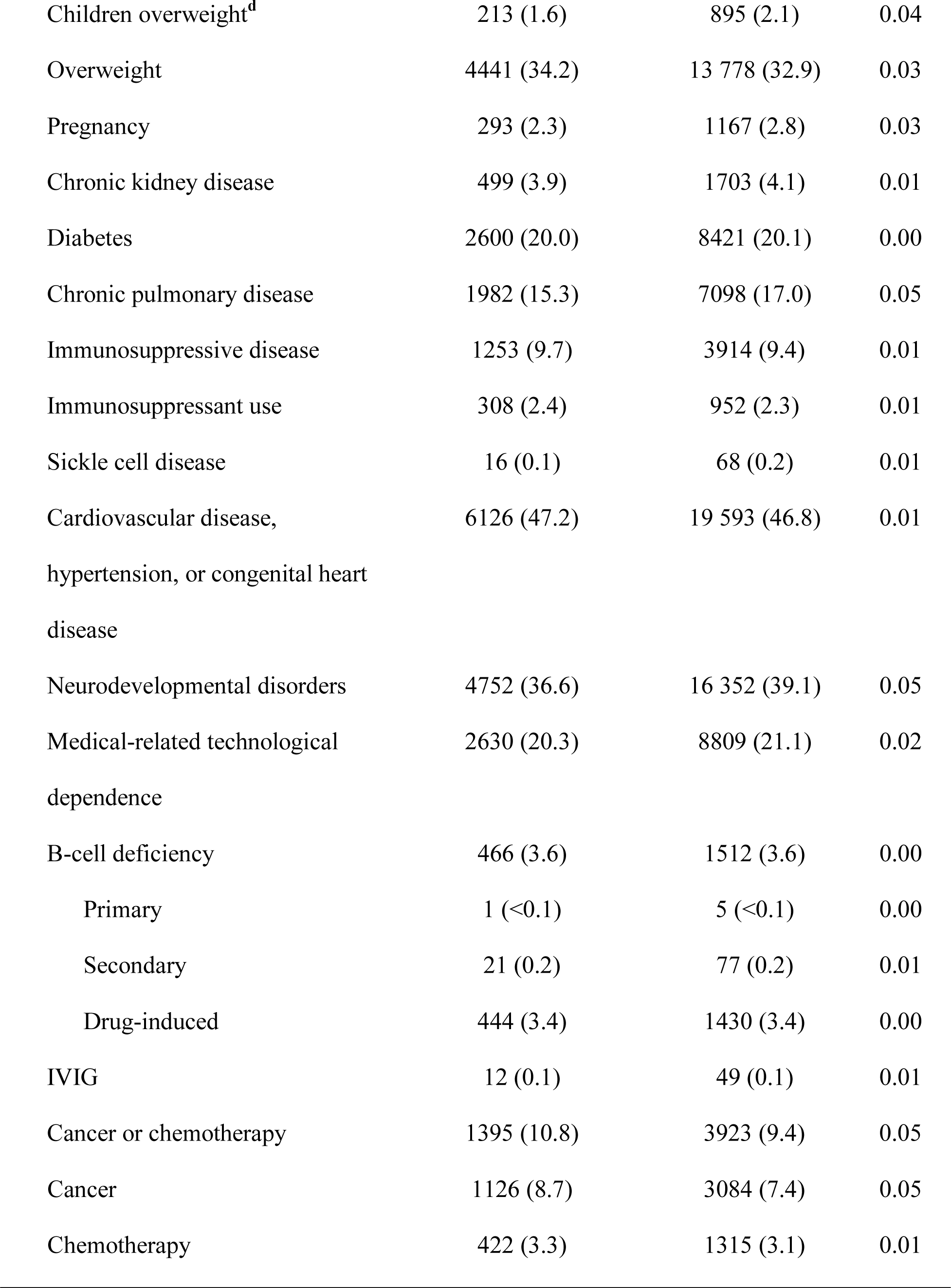
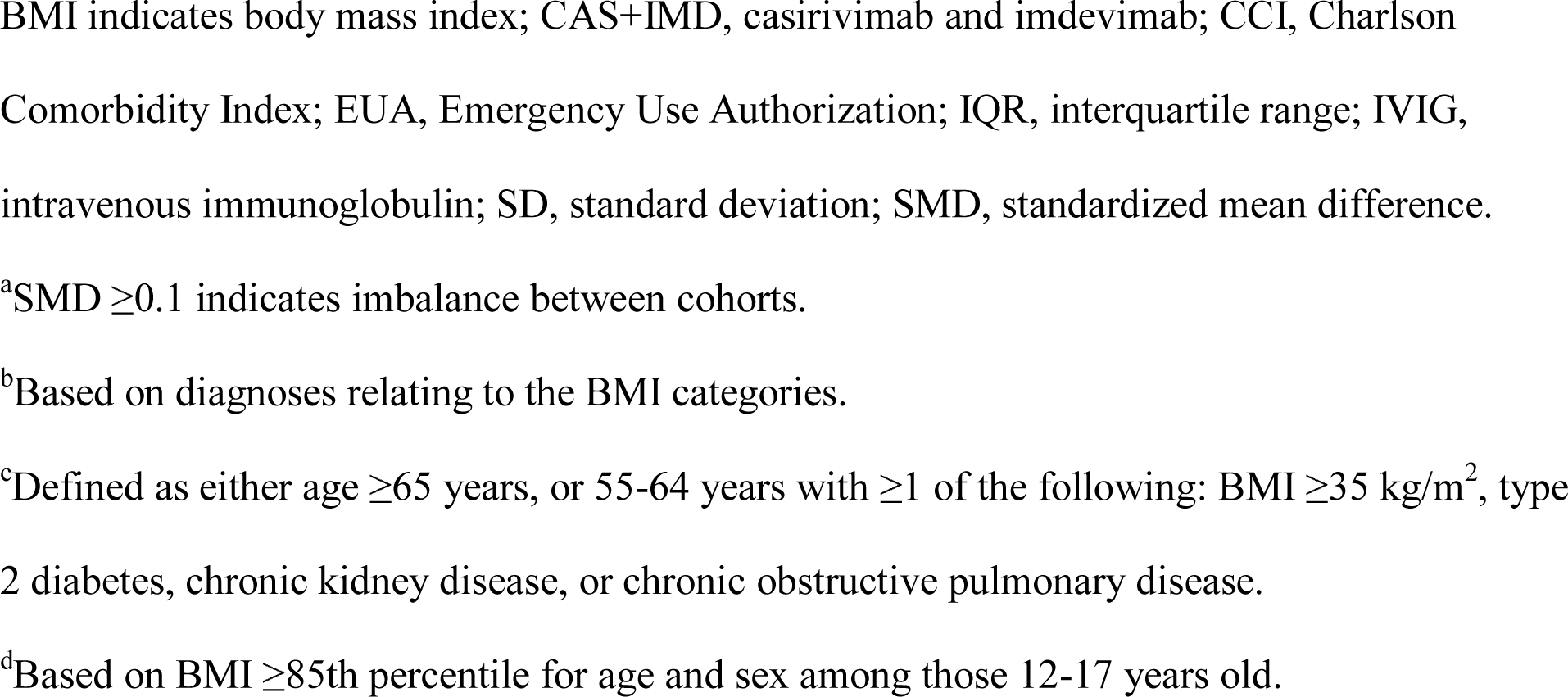
Baseline Characteristics of the Matched Cohorts.

### Primary analysis

The 30-day risk of the composite outcome was 1.9% (95% CI, 1.7-2.2) in the SC CAS+IMD-treated cohort (247 events) and 4.4% (95% CI, 4.2-4.6) in the EUA-eligible untreated cohort (1822 events) (Figure 1A). The 30-day mortality risk was lower in the treated cohort vs the untreated cohort: 0.1% (95% CI, 0.1-0.2; 11 deaths) and 0.3% (95% CI, 0.3-0.4; 128 deaths), respectively (Figure 1B).

In the adjusted model after applying the correction factor, treatment with SC CAS+IMD was associated with a 49% lower risk of the composite endpoint vs the untreated patients (aHR 0.51; 95% CI, 0.46-0.58) (Figure 2). Treatment was also associated with a 67% lower 30-day risk of all-cause mortality (aHR 0.33, 95% CI, 0.18-0.60) (Figure 2).

**Figure 2.**
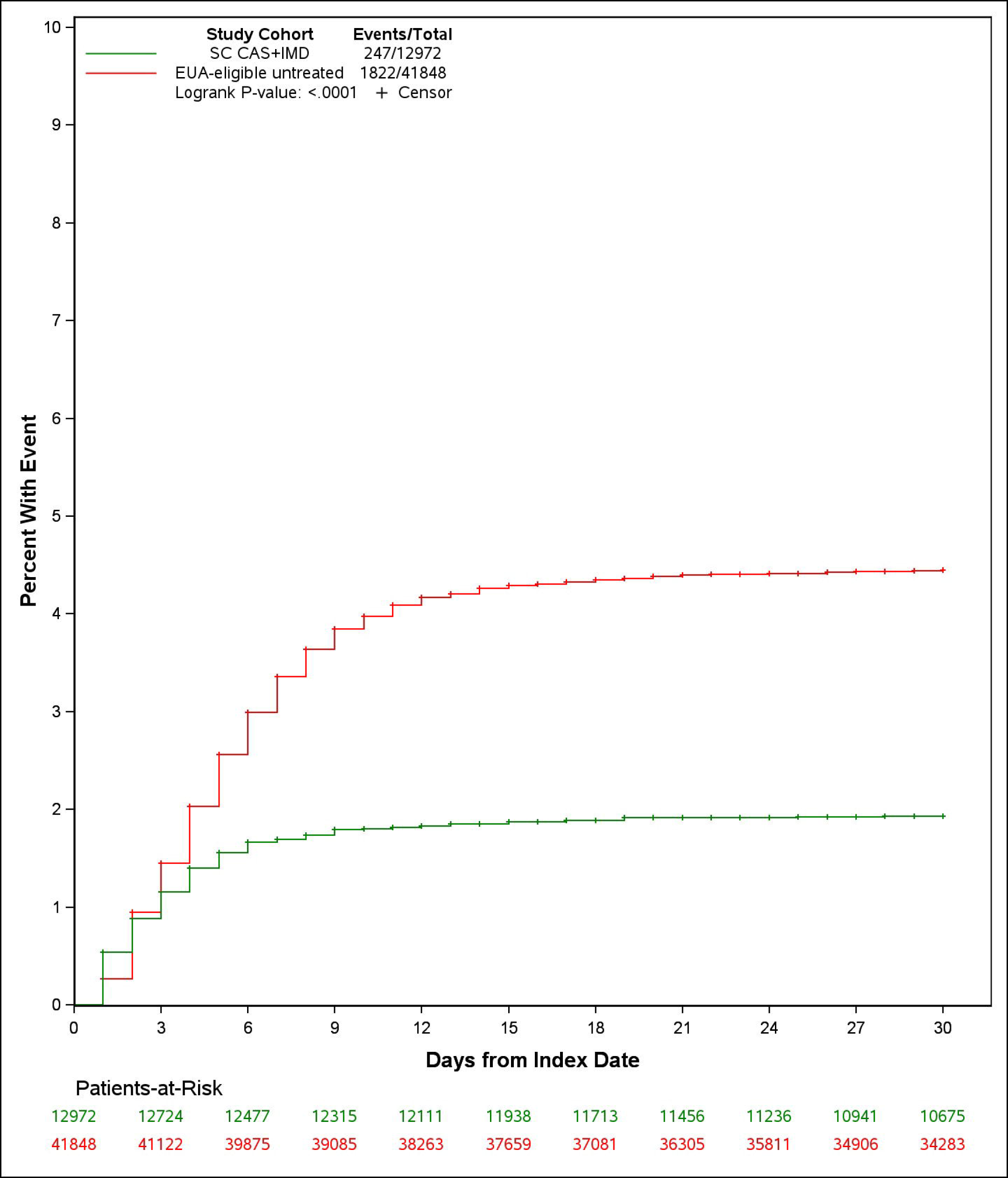
Adjusted Hazard Ratios of 30-Day All-cause Mortality or COVID-19-Related Hospitalization Among Patients Diagnosed With COVID-19 in the Outpatient Setting. Square size corresponds to the total sample available for analysis. aHR indicates adjusted hazard ratio; CAS+IMD, casirivimab and imdevimab; CI, confidence interval; EUA, Emergency Use Authorization; SC, subcutaneous.

The results of the sensitivity analyses were consistent; patients treated with SC CAS+IMD experienced 49-54% lower adjusted 30-day risk of mortality or COVID-19-related hospitalizations compared to EUA-eligible untreated patients (Figure 2).

### Subgroup analyses

The 30-day risk of the composite outcome generally increased with older age and was higher among those at elevated risk or who were immunocompromised (B-cell deficient) or unvaccinated relative to those without these risk factors (Table 2). The aHRs for treatment with SC CAS+IMD vs no treatment were generally consistent across subgroups (Figure 3). While the CIs were wide in the subgroups defined by age 12-17 years and secondary B-cell deficiency, the point estimates suggested that SC CAS+IMD treatment was beneficial. Results were inconclusive among patients with primary B-cell deficiencies due to a lack of observed events.

**Figure 3.**
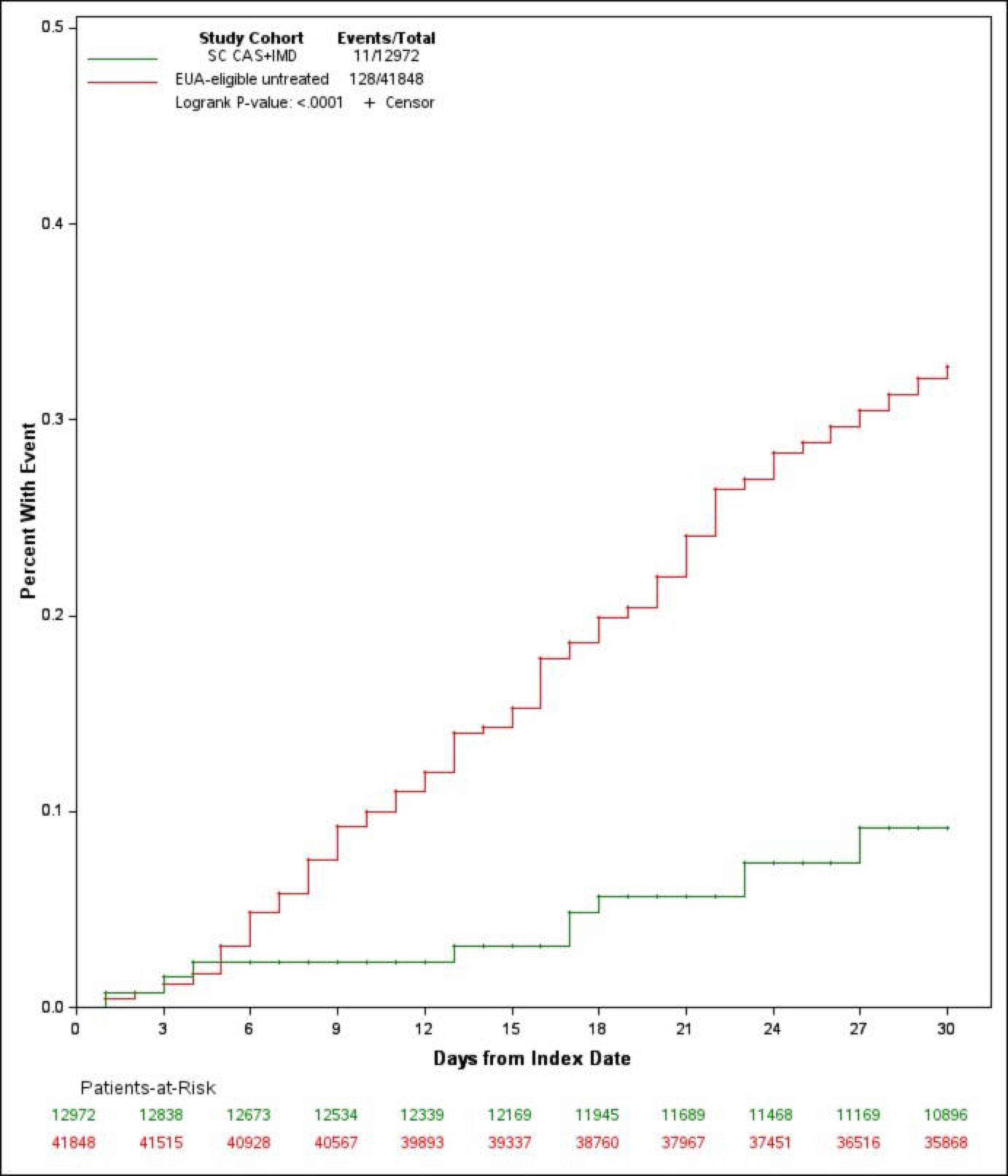
Adjusted Hazard Ratios of 30-Day All-cause Mortality or COVID-19-Related Hospitalization in Subgroups of Patients Diagnosed With COVID-19 in the Outpatient Setting. Square size corresponds to the total sample available for analysis. aHR indicates adjusted hazard ratio; CAS+IMD, casirivimab and imdevimab; CI, confidence interval; EUA, Emergency Use Authorization; N/A, not available; SC, subcutaneous. ^a^Elevated risk defined as age ≥65 years, or ≥55 years with body mass index ≥35, type 2 diabetes, chronic obstructive pulmonary disease, or chronic kidney disease.

**Table 2.**
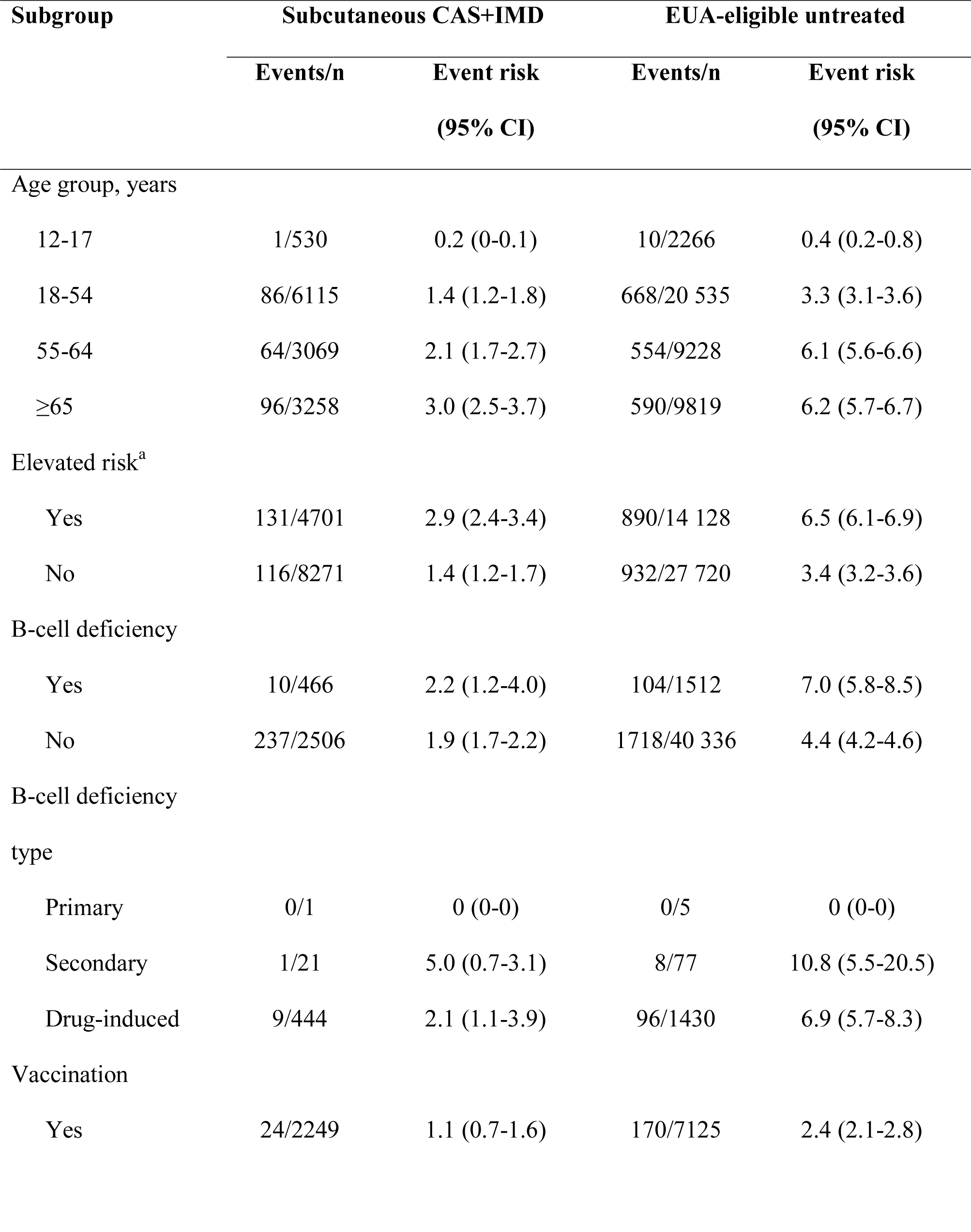
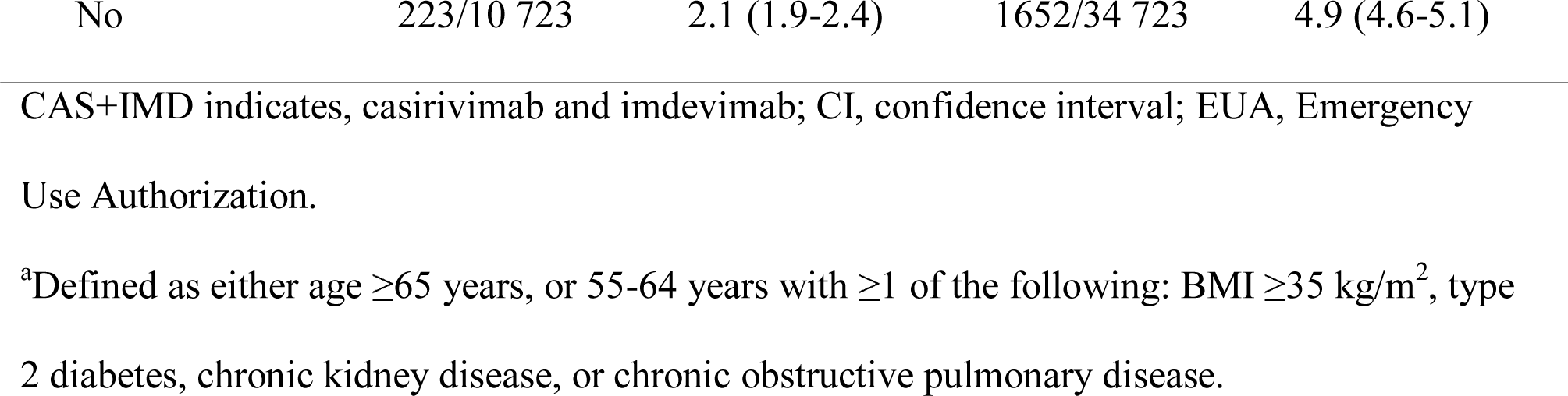
30-Day Risk of the Composite Event of All-Cause Mortality or COVID-19-Related Hospitalization in Subgroups

| Subgroup | Subcutaneous CAS+IMD |  | EUA-eligible untreated |  |
| --- | --- | --- | --- | --- |
|  | Events/n | Event risk<br>(95% CI) | Events/n | Event risk<br>(95% CI) |
| Age group, years |  |  |  |  |
| 12-17 | 1/530 | 0.2 (0-0.1) | 10/2266 | 0.4 (0.2-0.8) |
| 18-54 | 86/6115 | 1.4 (1.2-1.8) | 668/20 535 | 3.3 (3.1-3.6) |
| 55-64 | 64/3069 | 2.1 (1.7-2.7) | 554/9228 | 6.1 (5.6-6.6) |
| ≥65 | 96/3258 | 3.0 (2.5-3.7) | 590/9819 | 6.2 (5.7-6.7) |
| Elevated risk<sup>a</sup> |  |  |  |  |
| Yes | 131/4701 | 2.9 (2.4-3.4) | 890/14 128 | 6.5 (6.1-6.9) |
| No | 116/8271 | 1.4 (1.2-1.7) | 932/27 720 | 3.4 (3.2-3.6) |
| B-cell deficiency |  |  |  |  |
| Yes | 10/466 | 2.2 (1.2-4.0) | 104/1512 | 7.0 (5.8-8.5) |
| No | 237/2506 | 1.9 (1.7-2.2) | 1718/40 336 | 4.4 (4.2-4.6) |
| B-cell deficiency type |  |  |  |  |
| Primary | 0/1 | 0 (0-0) | 0/5 | 0 (0-0) |
| Secondary | 1/21 | 5.0 (0.7-3.1) | 8/77 | 10.8 (5.5-20.5) |
| Drug-induced | 9/444 | 2.1 (1.1-3.9) | 96/1430 | 6.9 (5.7-8.3) |
| Vaccination |  |  |  |  |
| Yes | 24/2249 | 1.1 (0.7-1.6) | 170/7125 | 2.4 (2.1-2.8) |
| No | 223/10 723 | 2.1 (1.9-2.4) | 1652/34 723 | 4.9 (4.6-5.1) |

CAS+IMD indicates, casirivimab and imdevimab; CI, confidence interval; EUA, Emergency
Use Authorization.
<sup>a</sup>Defined as either age $\geq 65$ years, or 55-64 years with $\geq 1$ of the following: BMI $\geq 35$ kg/m<sup>2</sup>, type 2 diabetes, chronic kidney disease, or chronic obstructive pulmonary disease.

## Discussion

This real-world study found that SC CAS+IMD is effective in the treatment of patients diagnosed with COVID-19 and managed in the outpatient setting,^24^ and further supports the benefits of treatment reported in clinical trials and smaller real-world studies.^2, 14, 15, 19, 23^ Importantly, this study was conducted during the Delta-dominant period, supporting the effectiveness of SC CAS+IMD against this variant. As expected in the EUA-eligible untreated control group, worse COVID-19 outcomes were observed that generally increased with age and were highest in those who were at elevated risk, immunocompromised (i.e., B-cell deficient) or unvaccinated. Treatment with SC CAS+IMD resulted in a 49% reduction in the risk of hospitalization/mortality compared with the EUA-eligible untreated patients and these findings were robust across sensitivity analyses. This study also demonstrated that the effect of SC CAS+IMD treatment was maintained across subgroups of patients at greater risk of poor COVID-19 outcomes.

The observed reduction is consistent with a 56% reduction in 28-day mortality/hospitalization reported in a previous study of 652 patients treated with SC CAS+IMD.^24^ While the hospitalization outcome of that study was for any cause, it also evaluated IV CAS+IMD and suggested no significant difference in outcomes between IV and SC administration. The results are also consistent with real-world effectiveness reported in claims-based analyses, including one that encompassed a period when the Delta variant was dominant, although SC administration was not specifically evaluated.^25, 29^

Results of the subgroup analyses are further concordant with the previous claims database studies;^25, 29^ the risk of outcomes was reduced among CAS+IMD-treated patients across all subgroups, demonstrating effectiveness regardless of age or vaccination status and in patients who were at elevated risk or immunocompromised. In particular, treatment effects in vaccinated patients were similar to those who were not vaccinated, suggesting the utility of therapy in EUA-eligible patients with breakthrough infections and in those who are unwilling to be vaccinated or for whom COVID-19 vaccines are less effective. Of clinical relevance is that the largest observed treatment effect was among patients who were immunocompromised, specifically those with secondary B-cell deficiencies, who are at increased risk for severe COVID-19 and poorer outcomes.^38, 39^ However, given the limited number of patients with primary B-cell deficiencies, the benefit of treatment could not be confirmed in this subgroup, although prior real-world studies have demonstrated that this patient subgroup also benefits from CAS+IMD treatment.^25, 29^

## Limitations

Limitations include that viral load and symptoms, which are indicative of COVID-19 severity and may be predictive of outcomes,^40–42^ were not captured. If untreated EUA-eligible patients had less severe disease, which may be why they are untreated, the lack of information on symptoms and confounders may have resulted in bias against CAS+IMD and thus underestimate the treatment effect. Another limitation regards how BMI was captured, since being overweight or obese is a strong risk factor for poor outcomes.^43–46^ Since categorization of BMI was based on ICD-10 codes and most patients did not have their BMI recorded using an ICD diagnosis code, the results may be subject to residual confounding. Residual confounding may also have arisen because vaccination status, a potentially important confounder, is under-captured in the Komodo Health data. Additionally, given that a correction factor was applied to derive unbiased aHRs, the crude risks between treated and untreated patients are not directly comparable. Furthermore, to estimate the correction factor for SC CAS+IMD-treated patients without a recorded COVID-19 diagnosis, it was assumed that 10 days had elapsed between symptom onset and treatment, which may have inflated the correction factor and resulted in underestimating the treatment effect.

While the study period did not overlap with emergence of the Omicron variant, CAS+IMD is not expected to be active against Omicron.^9^ Last, the untreated control group consisted of patients who were never treated with COVID-19 mAbs during the study period, potentially resulting in a healthier cohort of patients that could bias results against treatment.

## CONCLUSIONS

This study reports the real-world benefits of SC CAS+IMD among patients diagnosed with COVID-19 in the ambulatory setting during the Delta-dominant period. Overall, SC CAS+IMD was associated with a 49% reduction in the risk of COVID-19-related hospitalization/mortality compared with EUA-eligible untreated controls, with benefits that were generally maintained across subgroups, including vaccinated patients. Given the emergence of new variants of concern, continual monitoring and re-assessment of real-world effectiveness is integral to updating management strategies and identifying factors that can further improve patient outcomes and reduce pandemic transmission.

## Author Contributions

Drs Jalbert and Wei had full access to all of the data in the study and take responsibility for the integrity of the data and the accuracy of the data analysis. Concept and design: Jalbert, Hussein, Mastey, Sanchez, Murdock, Wei Acquisition, analysis, or interpretation of data: Jalbert, Hussein, Mastey, Sanchez, Murdock, Wang, Farinas, Bussey, Duart, Wei Drafting of the manuscript: All authors Statistical analysis: Wang, Wei Administrative, technical, or material support: Mastey, Weinreich, Hirshberg

## Data Availability

All data generated or analyzed during this study are included in this published article.

## Acknowledgements

Medical writing support was provided by E. Jay Bienen, PhD, and was funded by Regeneron Pharmaceuticals, Inc. Data programming and analytics support was provided by Wenqin Qiang and Dehua Kan from KMK Consulting Inc. and was funded by Regeneron Pharmaceuticals, Inc.

