## Supplementary material for "Effectiveness of Subcutaneous Casirivimab and Imdevimab Relative to no COVID-19 Antibody Treatment Among Patients Diagnosed With COVID-19 in the Ambulatory Setting"

**eFigure 1. Study Attrition.**

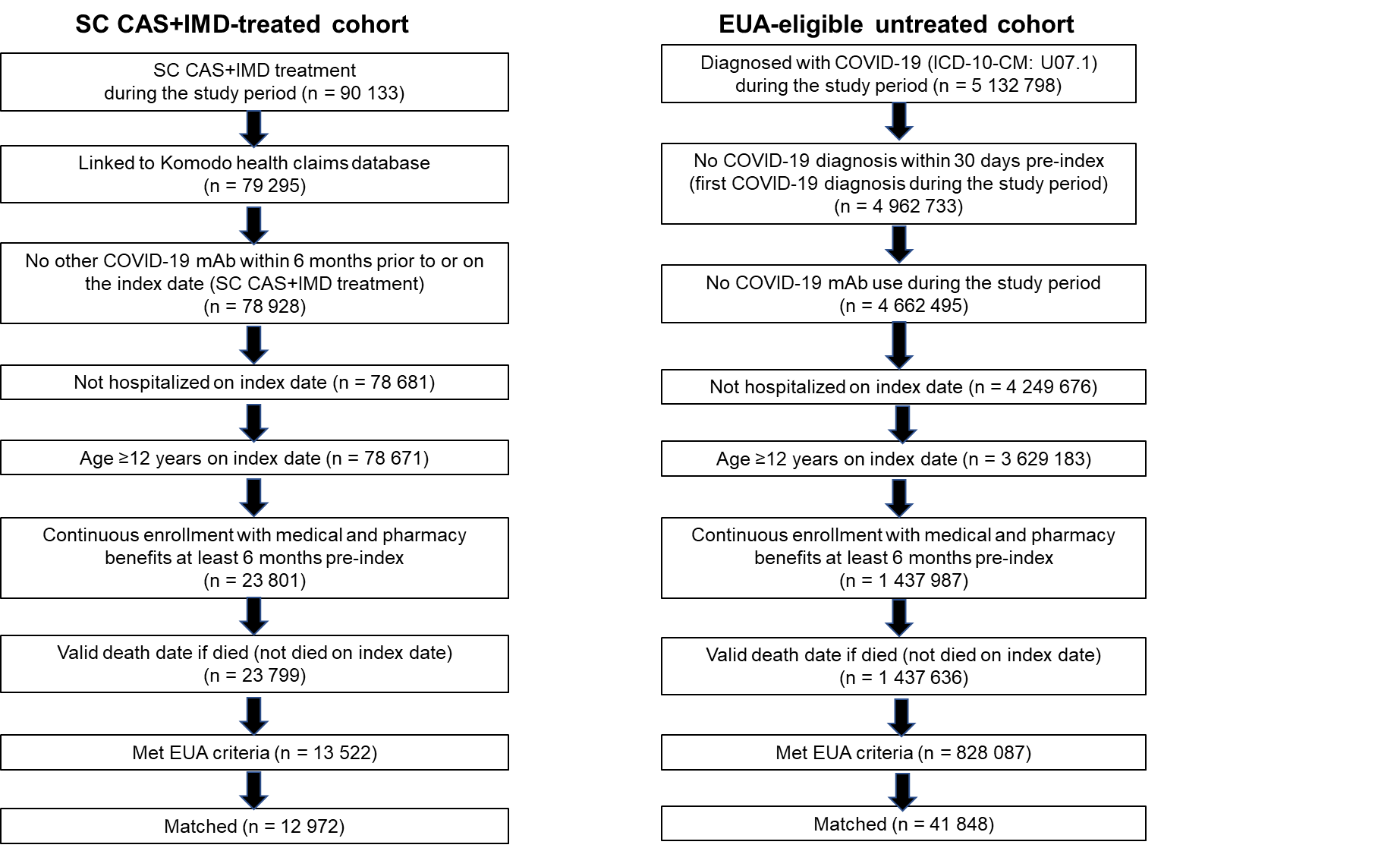

CAS+IMD indicates casirivimab and imdevimab; EUA, Emergency Use Authorization; mAb, monoclonal antibody; SC, subcutaneous

**eTable 1. B-Cell Deficiencies**

**Primary B-cell deficiencies (≥1 inpatient or ≥2 outpatient diagnoses ≥30 days apart during the baseline period, including index date)**

X-linked agammaglobulinemia

X-linked immunodeficiency with hyper-IgM

Selective IgA deficiency

Selective IgM deficiency

IgG subclass deficiency

Transient hypogammaglobulinemia of infancy

Common variable immunodeficiency (CVID)

Kappa/lambda light-chain deficiency

Severe combined immunodeficiency

Immunodysregulation polyendocrinopathyenteropathy X-linked syndrome (IPEX)

**Secondary causes of B-cell deficiency (≥1 inpatient or ≥2 outpatient diagnoses ≥30 days apart during the baseline period, including index date**

Multiple myeloma

Plasma cell leukemia

Acute lymphocytic leukemia (ALL)

Non-Hodgkin’s lymphoma (NHL)

Follicular lymphoma

Burkitt’s lymphoma

Diffuse B-cell lymphoma

Mantle cell lymphoma

Anaplastic large-cell lymphoma

Lymphoblastic lymphoma

Lymphoplasmacytic lymphoma

Marginal zone B-cell lymphoma/MALT lymphoma

Small-cell lymphocytic lymphoma

Non-Hodgkin’s lymphoma, unspecified/other

Hodgkin’s lymphoma (HL)

HIV/AIDS

**Drug-induced B-cell deficiencies (dispensation or administration within 3 months prior to index date)**

Rituximab

Ofatumumab (Kesimpta)

Ocrelizumab (Ocrevus)

Obinutuzumab

Inotuzumab ozogamicin

Blinatumomab

Alemtuzumab

Tocilizumab (Actemra)

Sarilumab (Kevzara)

Siltuximab

Belimumab (Benlysta)

Methotrexate

Mycophenolate mofetil (Cellcept, Myfortic)

Azathioprine (Imuran)

Systemic radiation therapy (excluding localized)

Chemotherapy

**eTable 2. Baseline Characteristics of the Unmatched Cohorts**

| **Variable** | **Subcutaneous CAS+IMD**  **(n = 13 522)** | **EUA-eligible untreated**  **(n = 828 087)** | **SMD^a^** |
| --- | --- | --- | --- |
| Age, years |  |  |  |
| Mean (SD) | 52.6 (17.0) | 44.8 (19.6) | 0.43 |
| Median (IQR) | 54 (24) | 44 (31) | — |
| Range | 12-88 | 12-88 | — |
| Age group, years, No. (%) |  |  |  |
| 12-17 | 555 (4.1) | 88 500 (10.7) | 0.25 |
| 18-34 | 1532 (11.3) | 191 623 (23.1) | 0.32 |
| 35-44 | 2024 (15.0) | 135 603 (16.4) | 0.04 |
| 45-54 | 2669 (19.7) | 132 127 (16.0) | 0.10 |
| 55-64 | 3248 (24.0) | 135 690 (16.4) | 0.19 |
| 65-74 | 2329 (17.2) | 85 358 (10.3) | 0.20 |
| 75-84 | 893 (6.6) | 40 532 (4.9) | 0.07 |
| ≥85 | 272 (2.0) | 18 654 (2.3) | 0.02 |
| Sex, No. (%) |  |  |  |
| Female | 7631 (56.4) | 511 799 (61.8) | -0.11 |
| Male | 5891 (43.5) | 316 287 (38.2) | -0.11 |
| Region, No. (%) |  |  |  |
| Midwest | 492 (3.6) | 170 747 (20.6) | 0.54 |
| Northeast | 664 (4.9) | 141 839 (17.1) | 0.40 |
| South | 11 983 (88.6) | 411 957 (49.8) | 0.93 |
| West | 383 (2.8) | 103 544 (12.5) | 0.37 |
| Florida resident, No. (%) | 11 029 (81.6) | 75 168 (9.1) | 2.12 |
| BMI category, No. (%)^b^ |  |  |  |
| Not overweight | 672 (5.0) | 30 744 (3.7) | 0.06 |
| Overweight (25 - < 30 kg/m^2^) | 1694 (12.5) | 64 061 (7.7) | 0.16 |
| Obese (30 - < 35 kg/m^2^) | 1414 (10.5) | 65 346 (7.9) | 0.09 |
| Severely obese (35 - < 40 kg/m^2^) | 747 (5.5) | 44 053 (5.3) | 0.01 |
| Morbidly obese (≥ 40 kg/m^2^) | 694 (5.1) | 55 988 (6.8) | 0.07 |
| Missing | 8301 (61.4) | 567 895 (68.6) | 0.15 |
| CCI score, mean (SD) | 1.01 (1.71) | 1.02 (1.76) | 0.01 |
| All-cause resource use during baseline period, No. (%) |  |  |  |
| Hospitalization | 1338 (9.9) | 94 769 (11.4) | 0.05 |
| Emergency room | 2724 (20.1) | 187 815 (22.7) | 0.06 |
| Month of index date, No. (%) |  |  |  |
| August 2021 | 3505 (25.9) | 404 710 (48.9) | 0.49 |
| September 2021 | 7650 (56.6) | 274 446 (33.1) | 0.48 |
| October 2021 | 2367 (17.5) | 148 931 (18.0) | 0.01 |
| Vaccinated, No. (%) | 2363 (17.5) | 131 169 (15.8) | 0.04 |
| Elevated risk, No. (%)**^c^** | 4978 (36.8) | 217 510 (26.3) | 0.23 |
| EUA criteria, No. (%) |  |  |  |
| Age ≥ 65 years | 3494 (25.8) | 144 544 (17.5) | 0.21 |
| Children overweight**^d^** | 221 (1.6) | 29 725 (3.6) | 0.12 |
| Overweight | 4549 (33.6) | 229 448 (27.7) | 0.13 |
| Pregnancy | 301 (2.2) | 45 265 (5.5) | 0.17 |
| Chronic kidney disease | 505 (3.7) | 30 714 (3.7) | 0.00 |
| Diabetes | 2662 (19.7) | 156 353 (18.9) | 0.02 |
| Chronic pulmonary disease | 2026 (15.0) | 168 500 (20.4) | 0.14 |
| Immunosuppressive disease | 1336 (9.9) | 68 705 (8.3) | 0.06 |
| Immunosuppressant use | 358 (2.7) | 16 735 (2.0) | 0.04 |
| Sickle cell disease | 198 (0.1) | 2132 (0.3) | 0.03 |
| Cardiovascular disease, hypertension, or congenital heart disease | 6299 (46.6) | 343 071 (41.4) | 0.10 |
| Neurodevelopmental disorders | 4865 (36.0) | 381 511 (46.1) | 0.21 |
| Medical-related technological dependence | 2697 (20.0) | 182 427 (22.0) | 0.05 |
| B-cell deficiency | 497 (3.7) | 27 482 (3.3) | 0.02 |
| Primary | 3 (<0.1) | 105 (<0.1) | 0.01 |
| Secondary | 22 (0.2) | 1356 (0.2) | 0.00 |
| Drug-induced | 472 (3.5) | 26 021 (3.2) | 0.02 |
| IVIG | 12 (0.1) | 740 (0.1) | 0.00 |
| Cancer or chemotherapy | 1450 (10.7) | 66 894 (8.1) | 0.09 |
| Cancer | 1167 (8.6) | 50 621 (6.1) | 0.10 |
| Chemotherapy | 444 (3.3) | 24 695 (3.0) | 0.02 |

BMI indicates body mass index; CAS+IMD, casirivimab and imdevimab; CCI, Charlson Comorbidity Index; EUA, Emergency Use Authorization; IQR, interquartile range; IVIG, intravenous immunoglobulin; SD, standard deviation; SMD, standardized mean difference.

^a^SMD ≥ 0.1 indicates significant imbalance between cohorts.

^b^Based on diagnoses relating to the BMI categories.

^c^Defined as either age ≥65 years, or 55-64 years with ≥1 of the following: BMI ≥35 kg/m^2^, type 2 diabetes, chronic kidney disease, or chronic obstructive pulmonary disease.

^d^Based on BMI ≥ 85th percentile for age and sex among those 12-17 years old.
